# A Quality Improvement Initiative: Improving Time-to-Antibiotics for Pediatric Oncology Patients with Fever and Suspected Neutropenia

**DOI:** 10.1101/2020.11.25.20233205

**Authors:** David E Kram, Kia Salafian, Sarah M Reel, Emily Nance Johnson, Brianna Borsheim, Thomas B Russell, Will A Pearsall, Michael Mitchell, Chad McCalla

**Affiliations:** Division of Pediatric Hematology and Oncology, Wake Forest School of Medicine, Winston Salem, NC, USA; Wake Forest School of Medicine, Winston Salem, NC, USA; Department of Pediatrics, University of Cincinnati School of Medicine, Cincinnati, OH, USA; Department of Biostatistics and Data Science, Wake Forest School of Medicine, Winston Salem, NC, USA; Pharmacy Department, Wake Forest School of Medicine, Winston Salem, NC, USA; Division of Pediatric Emergency Medicine, Wake Forest School of Medicine, Winston Salem, NC, USA

**Author notes:** **Corresponding Author:** David E Kram.

## Abstract

**Background:** There is a high risk for adverse outcomes in immunocompromised, neutropenic pediatric oncology patients with fever if antibiotics are not received in a timely manner. As the absolute neutrophil count is typically unknown at the onset of fever, rapid antibiotic administration for all pediatric oncology patients with fever and suspected neutropenia is critical.

**Local Problem:** Despite efforts over the years to meet the standard of time-to-antibiotic delivery to within 60 minutes of arrival, audits revealed a prolonged and wide-ranging time-to-antibiotics in our pediatric emergency department.

**Methods:** We conducted a quality improvement initiative to reduce the time to antibiotic delivery for this high risk patient population. The setting was a pediatric emergency department in an academic tertiary care hospital. We assembled a multidisciplinary team to apply quality improvement methods to understand the problem, implement interventions, and evaluate the outcomes.

**Interventions:** We targeted delays in patient triage, delays in antibiotic ordering, delays in antibiotic choice, and delays in bedside indwelling Port-a-Cath accessing procedure. Among other interventions, we instituted three unique measures: ceftriaxone was administered to all pediatric oncology patients with suspected neutropenia and fever; a system of ordering antibiotics that was driven by the ED pharmacist obtaining a verbal order from the ED attending; and a nurse-driven order set triggered by a unique triage category which empowered nurses to access a patient’s central line, draw and send specified blood work, and deliver an intravenous antibiotic, all potentially before an ED provider sees the patient.

**Results:** Over a sustained 3 year period of time, the percentage of febrile oncology patients with suspected neutropenia who met the target time-to-antibiotic delivery rose from 51% to 96%. The mean time-to-antibiotic delivery fell from 58 minutes in the pre-intervention period to 28 minutes in the post-intervention period.

**Conclusions:** The interventions implemented by the multidisciplinary team, using quality improvement methodology, successfully improved the percentage of febrile oncology patients receiving antibiotics within 60 minutes of arrival to a pediatric emergency department.

## INTRODUCTION

Pediatric patients undergoing chemotherapy treatment for cancer are at risk for severe infections due to chemotherapy-induced neutropenia, a frequent complication of chemotherapy which puts patients in an immunocompromised state, and the presence of central venous access devices (CVADs), which increase the risk for invasive bacterial infections^1-4^. The coincidence of fever and severe neutropenia, commonly called fever and neutropenia (F&N), in child with a CVAD undergoing chemotherapy increases the risk for a life-threatening emergency greater than in patients with non-neutropenic fever (NNF)^5 6^. Given that fever may be the only symptom of an underlying infection at presentation and the absolute neutrophil count (ANC) is typically unknown at the onset of the fever, rapid assessment of and intervention for all children with cancer who have fever and suspected neutropenia is critical.

Research and consensus guidelines on this patient population largely focus on pediatric cancer patients with F&N. When presenting to the emergency department (ED), guidelines suggest that these patients should promptly be evaluated for infection, which includes urgent initiation of broad-spectrum antibiotics^2^. Rapid antibiotic administration within 60 minutes reduces mortality for patients with F&N and is a generally accepted “benchmark” goal for time-to-antibiotics (TTA) from arrival to the hospital as a measure of quality-of-care^7-9^. However, given that the ANC is typically unknown at the time of initial presentation, both patients with F&N and NNF must be taken into account when strategizing the optimal approach for these high risk populations.

Despite efforts over the years to meet the TTA goal to within 60 minutes of arrival for pediatric cancer patients with F&N, an 8-month retrospective analysis of children who were evaluated in the ED at Brenner Children’s Hospital revealed that only 51.22% of children received antibiotics within 60 minutes, and the mean TTA was 95.71 minutes. To address all potential factors which may contribute to a prolonged TTA, a quality improvement (QI) initiative was undertaken in the ED at Brenner Children’s Hospital which aimed to improve the TTA in pediatric cancer patients with a CVAD and suspected neutropenia. This QI initiative aimed not only to achieve the benchmark goal of TTA within 60 minutes for all pediatric cancer patients with fever, but to succeed where similar previous QI initiatives fell short: sustainability for years following the expected acute period of heightened awareness and adherence to new QI protocols.

## METHODS

### Setting

The ED at Brenner Children’s Hospital is a 24-bed level 1 trauma pediatric ED which sees approximately 33,000 pediatrics patients every year. Brenner Children’s serves a large region which includes western North Carolina, parts of Virginia, South Carolina, and Tennessee, and also serves as a referral center for patients from farther away. The pediatric ED is open 24 hours per day, 7 days per week and is staffed by pediatric emergency medicine physicians.

Brenner Children’s Pediatric Oncology Program cares for approximately 150 patients on active treatment and with a CVAD in place. There is at least 1 pediatric hematologist/oncologist on call 24 hours a day.

### The Quality Improvement Team

A multidisciplinary quality improvement (QI) team was formed consisting of a pediatric oncology clinicians, pediatric ED physicians, pediatric oncology and pediatric ED nursing, ED registration staff, pediatric ED pharmacists, a patient/family advisor, and several members from the hospital’s quality and performance improvement (QPI) team.

### Planning the Interventions

The QI team used the Wake Forest Baptist Health QPI Model for Improvement (**Supplementary Fig. 1**), which is an adaptation of the Institute for Healthcare Improvement Model for Improvement that utilizes the Plan-Do-Study-Act cycle for QI. To address the AIMS, the QI team developed a Project Charter to establish the goals for this project, review the baseline TTA data from the previous 8 months, and hypothesize the problems with the current process. The fundamental goal of the project was a target TTA of 60 minutes for pediatric patients undergoing chemotherapy who have a CVAD and suspected neutropenia. The team then developed an “As-Is” Process Map (**Fig. 1**) to provide a detailed outline of the baseline process for treating pediatric patients with fever and suspected neutropenia in the pediatric ED. The process map was created during a team meeting that included the patient/family representative who validated the process based on their personal experiences, after which it was verified by QPI experts who went to the ED to observe the processes on multiple occasions. A Cause-and-Effect (Fishbone) Diagram (**Supplementary Fig. 2**) was created to identify key factors contributing to the delay in TTA within the baseline process. Based on the Cause-and-Effect Diagram, an Action Priority Matrix (**Supplementary Fig. 3**) was created to prioritize interventions based on the time each would require to implement and the expected impact each would have on the overall goal of decreasing TTA in this population of patients. Four points along the TTA process were identified which collectively contributed to delays, and included delays in patient triage, delays in antibiotic ordering, delays in antibiotic choice, and delays in bedside indwelling Port-a-Cath accessing procedure.

**Figure 1:**
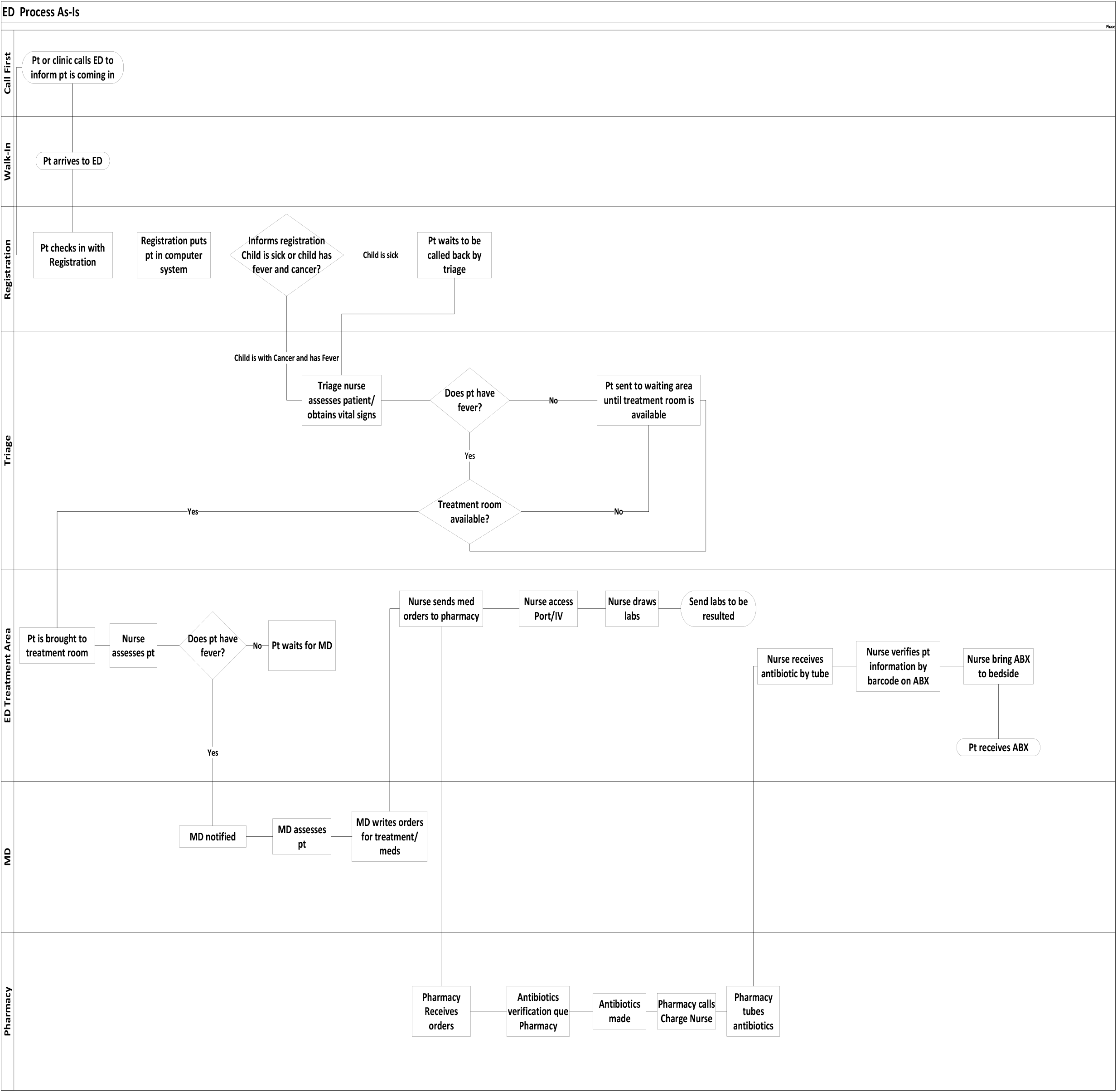
As-Is Process Map.

#### Delays in patient triage

Pediatric oncology patients with fever were not uniformly recognized as children with time-sensitive emergencies. To optimize the identification and triage assignment of this patient population, we adapted a “Fast Pass” (**Supplementary Fig. 4 and 5**) system used by other institutions that would signal to ED personnel the urgency of this encounter. Two copies were printed on durable plastic card and distributed to patients’ caregivers. Education on use was provided to both the caregivers and to all ED staff. This card thereafter triggered a chief complaint of “Oncology Fever” in the electronic health record (EHR), which triaged the patient as a high acuity. Importantly, signs and/or symptoms of shock in a presenting patient, whether or not they are a pediatric oncology patient with a Fast Pass, triggered the ED shock algorithm that would supersede this “Oncology Fever” triage acuity.

#### Delays in antibiotic ordering

Once a patient with fever and suspected neutropenia was roomed in the ED, there were delays in the ED clinician seeing and then ordering antibiotics for the patient, commonly due to expected fluctuations in ED patient volume and acuity. Thus there needed to be process whereby antibiotics could be administered to non-toxic patients without having to wait for a clinician to first see the patient. This issue was addressed with several interventions. A pediatric ED nursing order set was written and approved by the compliance committee at Brenner Children’s Hospital. Based on the presence of the accepted definition of fever in patient undergoing chemotherapy, the triage ED nurse would enter a set of orders in the EHR, which included orders to access the Port-a-Cath, obtain complete blood count with differential, and obtain one set of blood cultures from all central lines. Because nurses are not licensed to order antibiotics independently, our intervention then charged the triage nurse to call the ED pharmacist to notify them that the patient meets criteria for the Oncology Fever algorithm. This would trigger the ED pharmacist to call the supervising pediatric ED physician for a verbal order for antibiotics, which would enable the pharmacist to make and send to the bedside the antibiotic, and the nurse to administer it.

#### Delays in antibiotic choice

There was inconsistency and consequent delays in the antibiotic ordered. A previous QI effort to reduce TTA risk-stratified patients according to risk groups: high-risk patients were given ceftazidime (CFZ), which covers *pseudomonas aeruginosa* infections, and low-risk patients were given ceftriaxone (CTX). Over time, this led to a common practice of waiting for the ANC result before ordering an ANC level-specific antibiotic, which contributed to significant delays in TTA. Thus, we concluded that the antibiotic choice needed to be streamlined. While TTA has been shown to be a critical factor in pediatric patients with F&N, over half of emergency department presentations in the US for children with cancer and fever are for NNF^10^. Bacteremia is rare in pediatric patients with NNF, occurring in approximately 4-10% of febrile episodes, and outcomes for these patients who lack concerning clinical features are excellent.^6^ NNF is most commonly treated with CTX alone, which is a 3^rd^ generation cephalosporin that does not cover *pseudomonas aeruginosa* and is prone to bacterial resistence^11^. The rate of bacteremia in patients with F&N is higher, ranging from 16-25%, but bacteremia due to *pseudomonas aeruginosa* is quite rare, ranging from only 4-7% of cases with documented bacteremia^12-15^. Considering the very low risk for non-toxic appearing pediatric cancer patients with fever, with or without neutropenia, to have a CTX-insensitive or -resistant infection, and in collaboration with Brenner Children’s Pediatric Infectious Disease experts and hospital infection control, we decided to administer CTX (50 mg/kg, max 2 g) [or levofloxacin (10 mg/kg, max 750 mg) for cephalosporin-allergic patients] up-front to all non-toxic appearing pediatric cancer patients with fever and suspected neutropenia.

#### Delays in Port-a-Cath accessing

There were delays at various points during the bedside Port-a-Cath accessing procedure. The pediatric ED clinicians and nurses reported frequent patient-generated delays, including patients requesting a pediatric oncology nurse come to the ED access their Port-a-Cath or requesting to delay the Port-a-Cath access procedure until a local numbing agent had time to work (if they forgot to apply it themselves prior to arrival). Pediatric ED nurses also reported that, due to the typical turnover in nurse staffing, there was a wide range of nurse experience with the Port-a-Cath access procedure.

Finally, the patient/family advisors reported that the appropriate Port-a-Cath accessing needle size was not always easily and rapidly available. Our interventions focused on education: we began telling patients that the ED nurses would be the only nurses available to access the Port-a-Caths and that families should apply topical numbing agents to the Port-a-Cath prior to ED arrival because under no circumstances would topical numbing medication (other than instant freezing spray) be first applied in the ED. For the ED nursing staff, education was provided on an ongoing basis to capture new nurses regularly. Finally, information unique to each patient was added to the back of the “fast pass” card, which included the patient’s optimal Port-a-Cath needle size and preferred port dressing type (**Supplementary Fig. 5**). Each patient was also given one appropriately sized Port-a-Cath needle to bring with them to any ED encounter, in case the needed size was not readily available for any reason.

### Planning the Study of the Intervention

The measure selected for this initiative included comparisons between the pre-intervention and post-intervention periods for the primary outcome, the proportion of patients receiving antibiotics within 60 minutes of ED arrival, and the secondary outcome, the mean and median TTA. An unpowered exploratory outcome measured adverse events associated with administration of CTX as the first-line antibiotic for all patients (except cephalosporin-allergic patients). The population of interest was all pediatric oncology patients with a CVAD who presented to Brenner ED with fever (≥ 38.3°C once or ≥38.0°C twice in one hour) and suspected neutropenia. Suspected neutropenia was defined as the absence of an ANC value < 500 within the preceding 48 hours; an ANC < 500 within 48 hours was considered known neutropenia. IRB approval for this project was obtained, retrospective chart reviews were conducted, and data was entered into RedCap. As these interventions were made for the purpose of QI, consent was not obtained.

### Measures and Analysis

Descriptive statistics were calculated for all patient encounters and compared between the pre- and post-intervention groups. Encounters were considered individual observations, and patients could have been counted more than once in either or both of the groups if they had received antibiotics in the ED multiple times during the study time period. Categorical variables were summarized with counts and percentages, and comparisons were made using Chi-square or Fisher’s exact tests. Patients’ age at presentation was summarized with the mean, standard deviation, and range, and a T-test was used to compare groups. TTA was summarized with the median and range, and a Wilcoxon rank-sum test was used to compare groups. A scatterplot was created to show the observed times from arrival to antibiotic for the pre- and post-intervention time periods. Reference lines were added to indicate the implementation of the intervention and the goal TTA of 60 minutes. A frequency table shows number of encounters where the time to antibiotic was less than or greater than 60 minutes for both the pre- and post-intervention groups. All analyses were performed using SAS software, version 9.4 (SAS Institute, Cary, NC, USA).

## RESULTS

Over the course of this intervention, from July 1, 2017 through June 30, 2020, 360 patient encounters to the Brenner ED for fever and suspected neutropenia met criteria. The patients recorded in the pre-intervention period, from October 1, 2016 through June 30, 2017 (n = 41) were similar to those in the post-intervention period in terms of age, oncologic diagnosis, ethnicity, race, type of central access device, and time of arrival to the Emergency Department (**Table 1**). Both cohorts had a predominance of male patients. For the primary TTA outcome, the percentage of patients receiving antibiotics within one hour improved significantly from 51% to 96% (**Table 2, Fig. 2**). The time from arrival to time antibiotics administered decreased by 68%, from a mean of 96 minutes at baseline to 31 minutes post-intervention (**Table 1**). Similarly, the median time from arrival to antibiotics administered also decreased significantly by 52%, from a median time of 58 minutes at baseline to 28 minutes post-intervention, showing a significant decrease in the variation of TTA (**Table 1**).

**Figure 2:**
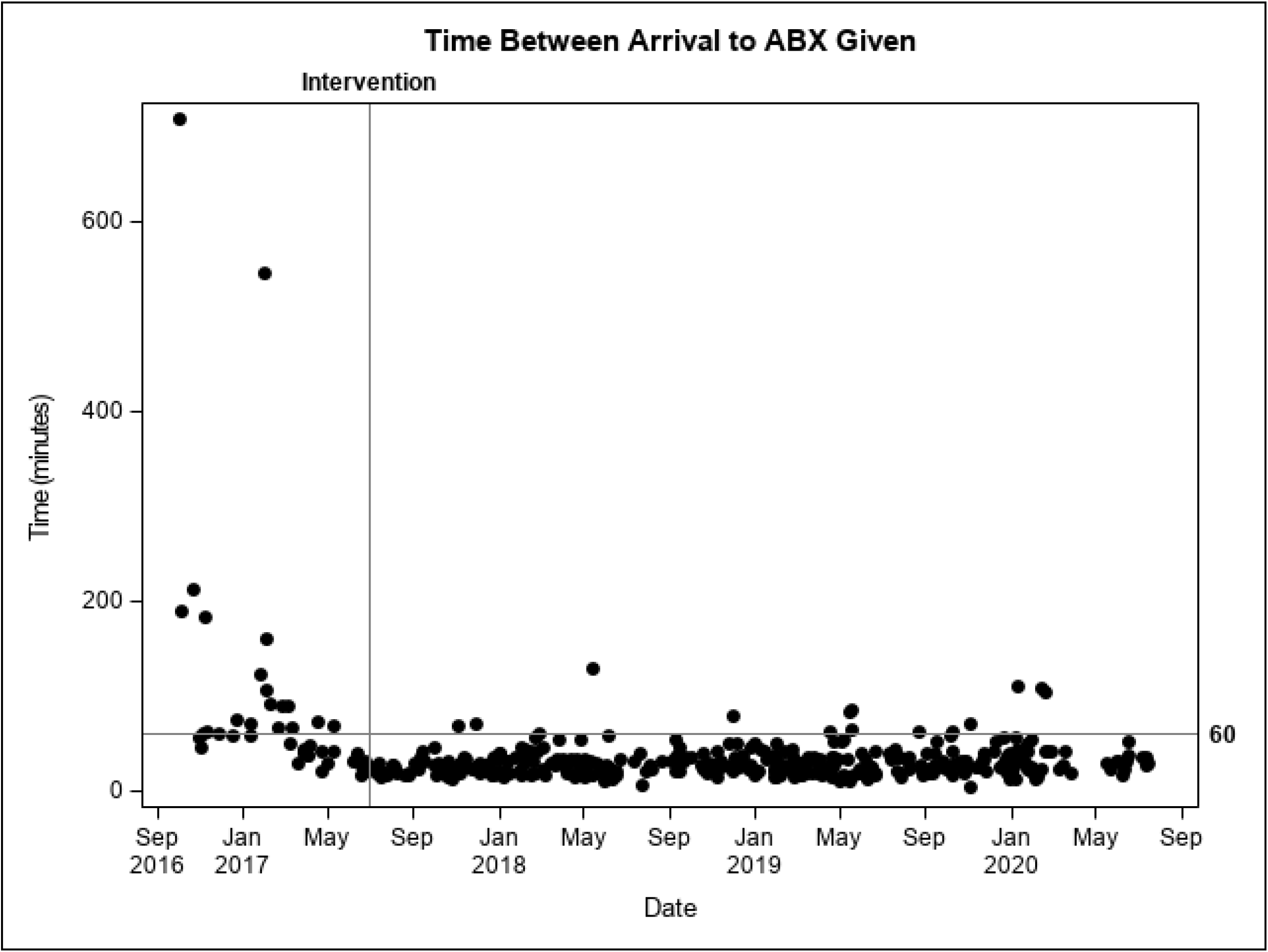
Time Plot including TTA (y axis) and date (x axis). The intervention time is labeled.

**Table 1:** Patient characteristics.

| Patients' Characteristics | Preintervention<br>(October 2016 – June 2017) | Postintervention<br>(July 2017 – June 2020) | P value |
| --- | --- | --- | --- |
| *N* | 41 | 360 |  |
| Mean age at presentation, *mean* (SD, range) | 7.03 (4.97, 0-19) | 6.96 (4.81, 0-21) | 0.9281*** |
| Sex, *n* (%) |  |  | 0.1233* |
| Male | 23 (56%) | 245 (68%) |  |
| Female | 18 (44%) | 115 (32%) |  |
| Oncologic Diagnosis, *n* (%) |  |  | 0.0655* |
| Hematologic malignancy | 25 (61%) | 268 (74%) |  |
| Non-hematologic malignancy | 16 (39%) | 92 (26%) |  |
| Ethnicity, *n* (%) |  |  | 0.8344* |
| Hispanic or Latino | 9 (22%) | 74 (21%) |  |
| Not Hispanic or Latino | 32 (78%) | 286 (79%) |  |
| Race, *n* (%) |  |  | 0.9887* |
| Asian | 1 (2%) | 15 (4%) |  |
| Black or African American | 4 (10%) | 36 (10%) |  |
| White | 26 (63%) | 230 (64%) |  |
| Other | 10 (24%) | 79 (22%) |  |
| Access, *n* (%) |  |  | 0.4804** |
| Port-a-Cath | 38 (93%) | 343 (95%) |  |
| PICC | 0 (0%) | 2 (1%) |  |
| CVC (Hickman catheter) | 2 (5%) | 12 (3%) |  |
| Peripheral IV | 1 (2%) | 3 (1%) |  |
| Time of arrival to the Emergency Department, *n* (%) |  |  | 0.4560* |
| 8 AM – 4 PM | 12 (29%) | 137 (38%) |  |
| 4 PM – 12 AM | 21 (51%) | 172 (48%) |  |
| 12 AM – 8 AM | 8 (20%) | 51 (14%) |  |
| Mean time from arrival to antibiotic given in minutes, *mean* (range); *median* | 95.71 (17-709)<br>58 | 30.61 (5-130)<br>28 | <0.0001**** |

\*Chi-square test
\*\*Fisher's Exact test
\*\*\*T-test
\*\*\*\*Wilcoxin rank-sum test

**Table 2:**
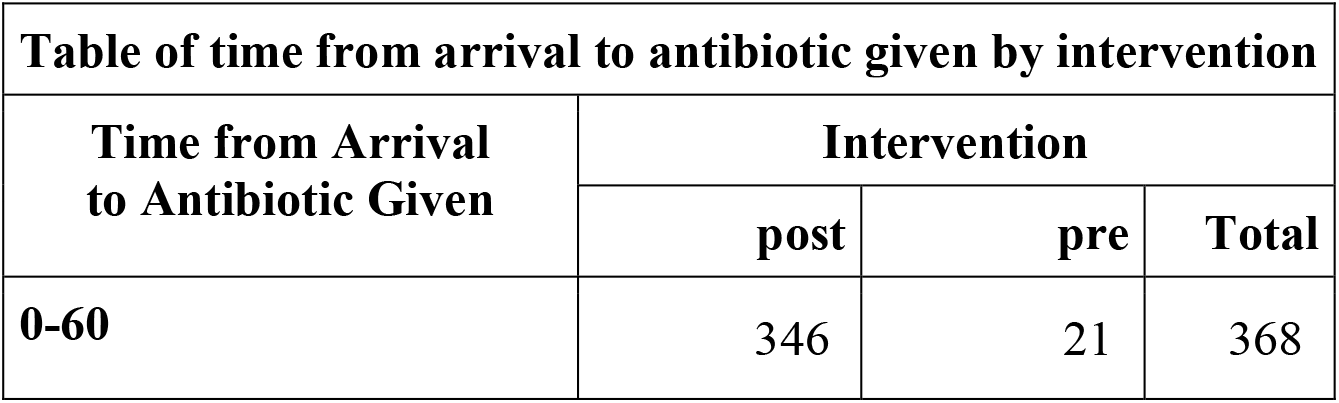
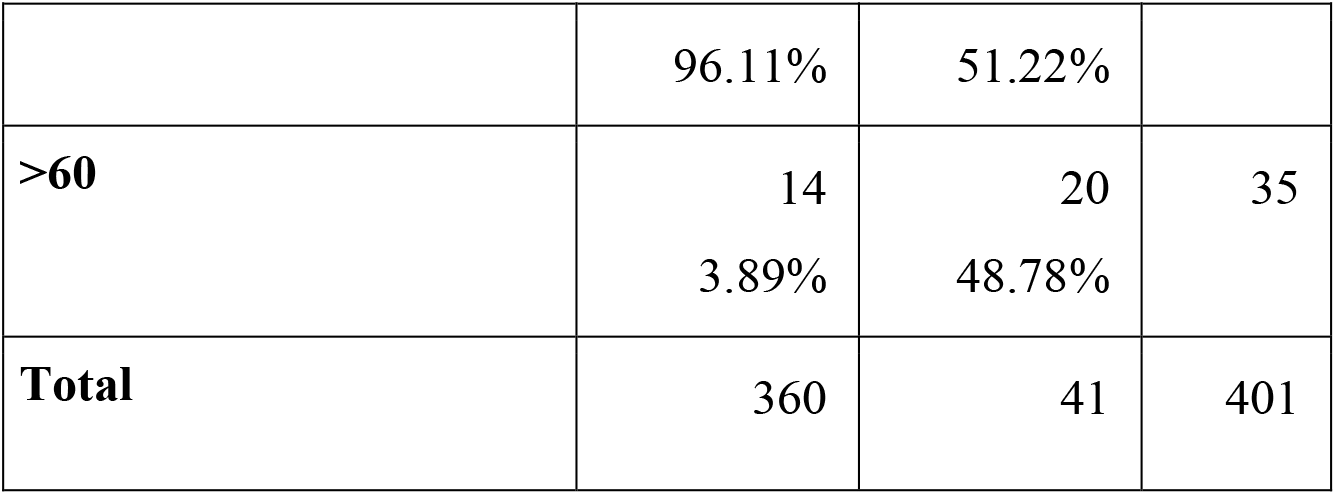
Time from arrival to antibiotic given by intervention period.

During the post-intervention period, there were 14 cases (3.89%) where patients received antibiotics after the 60-minute benchmark. The majority of these cases were due to difficulty accessing patients’ Port-a-Caths while in the ED; however, these patients were appropriately triaged in an urgent manner and recognized as pediatric oncology patients with fever. Twelve of the 14 patients received CTX during their stay and the majority of these patients did not require admission to the Pediatric Hematology Oncology unit nor further treatment. Of the two patients that did not receive CTX, one received levofloxacin due to a prior reaction to CTX. The other patient received CFZ as they were strongly believed to be neutropenic from a prior lab draw revealing a decreasing ANC greater than 48 hours prior. Overall, none of the patients whose antibiotic administration was delayed had adverse events or clinical worsening as a result of the delay. Of note, blood cultures from these patients did not grow any organisms.

As this was a QI initiative, where multiple interventions were made simultaneously, an understanding of the controlled impact of each intervention was not possible. Broadly, however, we evaluated the TTA in two discrete bins: the time from patient arrival to the ED until the time of antibiotic order entry into the EHR, and the time from antibiotic order entry into the EHR until the time of antibiotic administration to the patient. The first bin would potentially be impacted by the majority of our QI interventions, including the “Fast Pass,” implementing the nursing order set, and simplifying the antibiotic choice process. The second bin would potentially be impacted by ongoing education for ED nurses on Port-a-Cath accessing, the “Fast Pass” Port-a-Cath needle size information, and the availability of an appropriately sized needle. Other steps during this period, including antibiotic preparation by pharmacy, antibiotic tubing from the pharmacy to the bedside, the ED nurse collecting the antibiotic from the tube and bringing it to the bedside, or the ED nurse actually administering the antibiotic, were not specifically addressed by this QI project. The time from patient arrival to the ED until the time of antibiotic order entry into the EHR was significantly reduced (**Supplemental Table 1**). In an evaluable cohort of pre-intervention patients (n = 21), the time from arrival to antibiotic order entry into the EHR was 20.14 minutes compared to 11.40 minutes in the post-intervention cohort, representing a 44% reduction in time [pre-intervention = 20.14 minutes (range 7-66 minutes); post-intervention = 11.40 minutes (range 0-56 minutes), *p* = 0.0006)]. Together, these improvements simplified, streamlined, and made more rapid the processes of identifying pediatric oncology patients with fever, triaging and rooming them, accessing their Port-a-Caths, and entering a CTX order for the majority of patients. In contrast, the time from antibiotic order entry into the EHR until the antibiotic was actually administered to the patient was not statistically different in the pre- and post-intervention groups (pre-intervention = 22.43 minutes (range 5-65 minutes); post-intervention = 19.23 minutes (range 1-126 minutes), *p* = 0.06).

Of the 360 patient encounters that took place in the post-intervention period, 14 patients (14/360, 3.9%) had positive blood cultures at presentation and outcome data was compiled. We assessed, in a non-powered approach, the safety of our approach to administer CTX up-front to all patients with fever and suspected neutropenia. Three patients (3/360, 0.8%) had blood cultures with multiple organisms grown, requiring multiple antibiotics to treat, including neither CTX nor CFZ; hence neither CTX nor CFZ would have been sufficient. Two patients (2/360, 0.6%) had bacteremia from an organism that would have been sensitive to CFZ, but was not sensitive to CTX. One patient (patient #2 in **Table 3**) was treated in the ED for fever, and was discharged home in good condition that day. The following day, when the blood culture grew an organism, the patient was brought back to the ED, given a second dose of CTX after repeat cultures, after which the patient developed septic shock and was admitted to the PICU on meropenem. The second patient (patient #10 in **Table 3**) had bacteremia with multiple organisms, so CFZ would not necessarily have been superior for this patient compared to CTX. Six patients (6/360, 1.7%) had infections that were sensitive to and treated with CTX; and five patients (5/360, 1.4%) had infections that were not sensitive to either CTX or CFZ. One patient (1/360, 0.3%; patient #11 in **Table 3**) had bacteremia from two organisms, one of which was sensitive to and treated with CTX, and one that was not sensitive to either CTX or CFZ. Overall, there appeared to be no adverse events directly as a consequence of using CTX upfront rather than a broader-spectrum, antipseudomonal agent and the majority of patients had hospital stays less than 10 days in length.

**Table 3:**
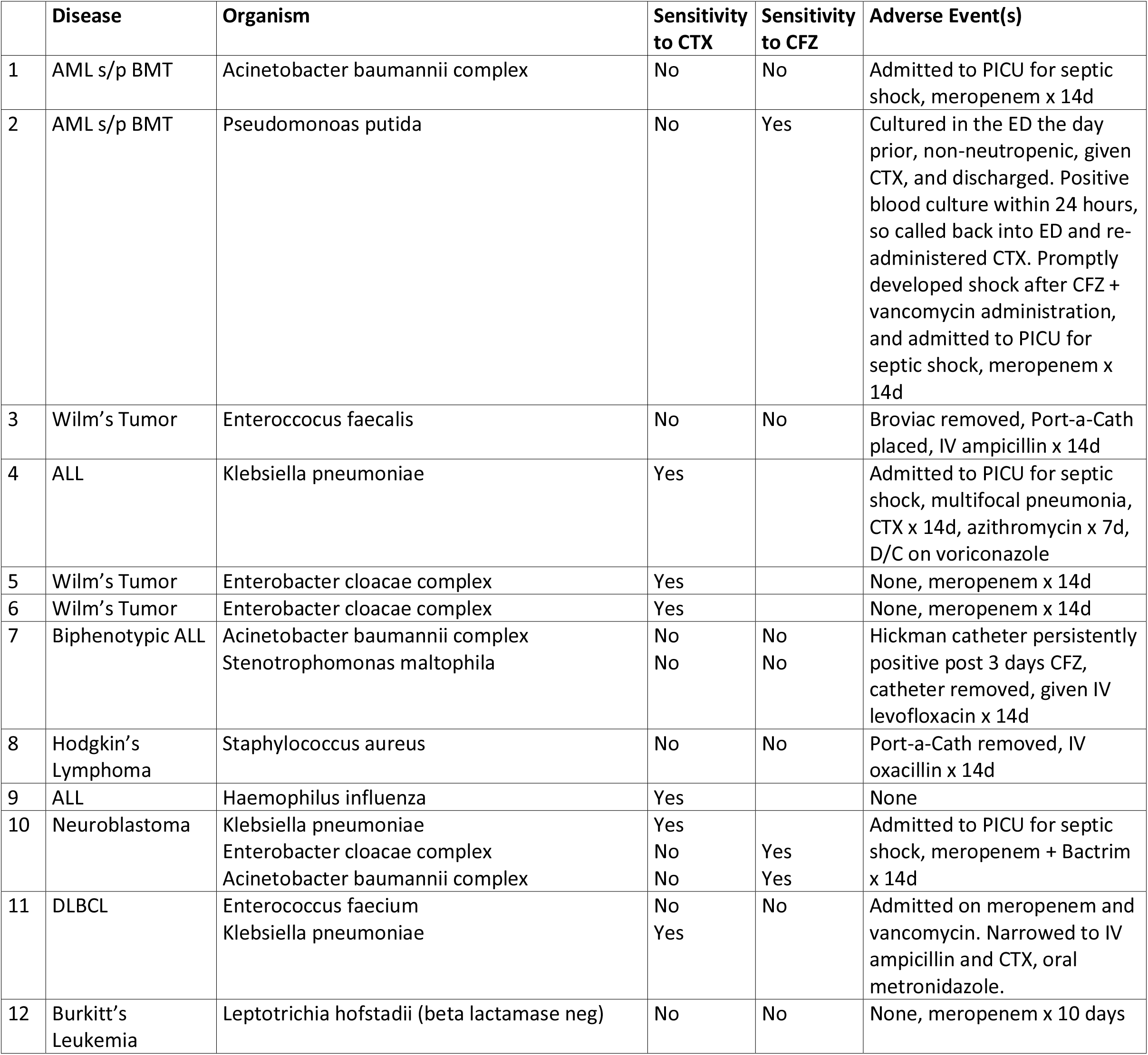
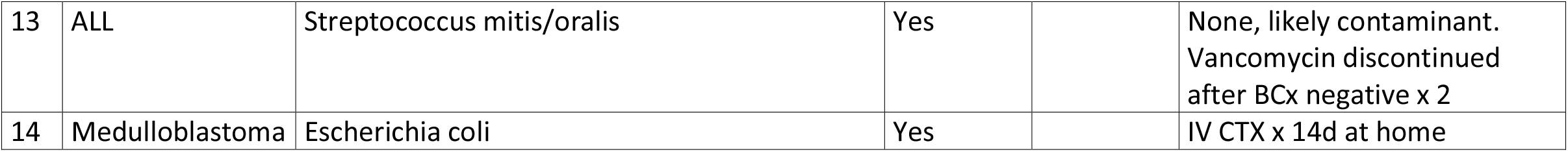
Patients with positive blood cultures, demographics, organisms, antibiotic sensitivities, and outcomes. ALL, acute lymphoblastic leukemia; AML, acute myeloid leukemia; BCx, blood culture; BMT, bone marrow transplant; DLBCL, diffuse large B cell lymphoma; D/C, discharge; IV, intravenous; PICU, pediatric intensive care unit.

## DISCUSSION

We successfully used QI methodology to significantly improve the percentage of pediatric oncology patients with fever and suspected neutropenia presenting to our pediatric ED receiving antibiotics within one hour from presentation from 54% to 96%. The collective interventions reduced the mean TTA significantly from 96 minutes pre-intervention to 31 minutes post-intervention. Importantly, this improvement has proved to be sustainable, maintaining this high rate of success over a period of three years. The use of a multidisciplinary team, and targeted efforts based off of the Process Map and Cause-and-Effect Diagram, enabled this project to be successful.

Our study included several similar interventions previously described in other TTA QI initiatives, including addressing triage issues with an intervention like a “Fast Pass,” optimizing the Port-a-Cath access procedure through education and providing an additional appropriately sized needle to the patient, and optimizing timeliness of antibiotic ordering and review by ED pharmacist^5 16 17^. These interventions were pivotal to our ultimate success. However, our QI initiative took several novel approaches, including the addition of a standing nursing order set, which empowered nurses to release blood work orders and CVAD accessing orders based on an appropriate “Oncology Fever” triage, and enabling the ED pharmacist to call the ED attending to obtain a verbal order for the antibiotic. These additions facilitated the reduced mean time from patient arrival to antibiotic ordering.

One additional unique feature of our intervention was the decision to administer ceftriaxone up-front to all pediatric oncology patients with fever and suspected neutropenia, an approach that has not been reported. The urge to distinguish and provide separate practice guidelines for patients with fever and neutropenia and patients with NNF is challenging because clinicians most often do not know whether the pediatric cancer patient with fever facing them is neutropenic or not^2 6 18^. Thus, an all-encompassing approach to pediatric oncology patients with fever and suspected neutropenia is crucial, and one that was successfully instituted and maintained by this QI initiative. Guidelines recommend *pseudomonas-*covering antibiotics^2^, such as CFZ, for children with fever and neutropenia, whereas multiple studies support empiric CTX for children with NNF^18-23^. In our experience, these disparate practices led to an overly complicated risk-assessment algorithm which ultimately contributed to delays in antibiotic administration. Knowing that children who were septic or toxic-appearing would be captured by the ED’s septic shock algorithm, thus mandating those patient received broader-than-CTX antimicrobials, and children with known neutropenia would receive appropriately broad antibiotics, we proceeded to use CTX for all remaining patients. In our unpowered review, we conclude that the practice of CTX up-front is safe and did not directly contribute to any adverse outcomes.

Several patients without known neutropenia and who were not treated for sepsis also received CFZ for various other reasons that we could not account for; however, our QI intervention acknowledged the importance of physician autonomy, thus enabling ED clinicians to ultimately choose the antibiotic for pediatric cancer patients with fever. In patients who received initial CTX, the positive blood culture rate was 3.5%. In contrast, in patients who did not receive upfront CTX (instead receiving CFZ, other antibiotic, or no antibiotic), the positive blood culture rate was 6.1%. This suggests that, for various reasons, those who received CFZ may have appeared more ill than the cohort who received CTX, thus further supporting our practice of initial CTX for all except those with sepsis, known neutropenia, or in whom the treating clinician chooses a different therapy.

Overall, multiple and simultaneous interventions were implemented in this study to affect rapid change. While we cannot identify the impact by each isolated intervention, the collective interventions certainly impacted the time from arrival to antibiotic order entry into the EHR, whereas the time from antibiotic order entry into the EHR until the antibiotic was administered to the patient was not impacted by our interventions. However, given our end results, a sustainably improved TTA within 60 minutes of ED presentation, we would argue that the time from arrival to antibiotic order entry into the EHR is the crucial time period. Future projects and other centers should focus on processes during this pivotal time period to affect change.

### Limitations

Our study has a number of limitations. The study was retrospective and uncontrolled, and thus subject to confounders, along with the inherit source of misinformation or errors associated with EHR documentation. This study was limited to patients within our hospital system, hence results may not be generalizable to all pediatric oncology populations elsewhere. Bacterial susceptibility certainly varies from city to city, hospital to hospital, and the practice of CTX for all patients with fever and suspected neutropenia may not be safe or feasible elsewhere. Finally, because all interventions in this project were implemented simultaneously, interventions could have unequally contributed to decreasing the mean TTA.

## CONCLUSION

Our study is similar to other QI initiatives aiming at improving TTA in a pediatric ED for pediatric oncology patients with fever. However, our QI initiative took several novel approaches, which are certainly replicable and should be considered by other centers facing the same TTA challenges. Working as a multidisciplinary team, including physicians, advanced practice practitioners, and nurses from both the pediatric ED and the pediatric oncology teams, ED registration staff, pediatric ED pharmacists, a patient/family advisor, and several members from the hospital’s quality and performance improvement (QPI) team, and our adherence to well-established QI principles and practices, we established sustained practice changes to this critical and common clinical scenario.

## Data Availability

Data is housed by Wake Forest School of Medicine Clinical and Translational Science Institute's REDCap platform and is available by request and with permission by David E Kram, MD (ORIC identifier https://orcid.org/0000-0002-1928-001X).

## ACKNOWLEDGMENTS

We are grateful to the pediatric emergency medicine providers at Brenner Children’s Hospital for allowing our QI team to work closely with them. We are also grateful to the QPI team who provided key expertise in the development, implementation, and tracking of this QI project.

## Notes

### Competing Interest Statement

The authors have declared no competing interest.

### Funding Statement

No external funding was received.

### Author Declarations

IRB approval for this project was obtained by Wake Forest University Health Sciences IRB (IRB #IRB00060779). As this was a QI retrospective chart review with data entry into RedCap, the study was deemed to be exempt from consent, and thus consent was not obtained.

