## Supplemental Figures 1-5, and Supplemental Table 1 for "A Quality Improvement Initiative: Improving Time-to-Antibiotics for Pediatric Oncology Patients with Fever and Suspected Neutropenia"

Supplementary Material

**Supplementary Figure 1. Wake Forest Baptist Health Performance Improvement Model for Improvement**

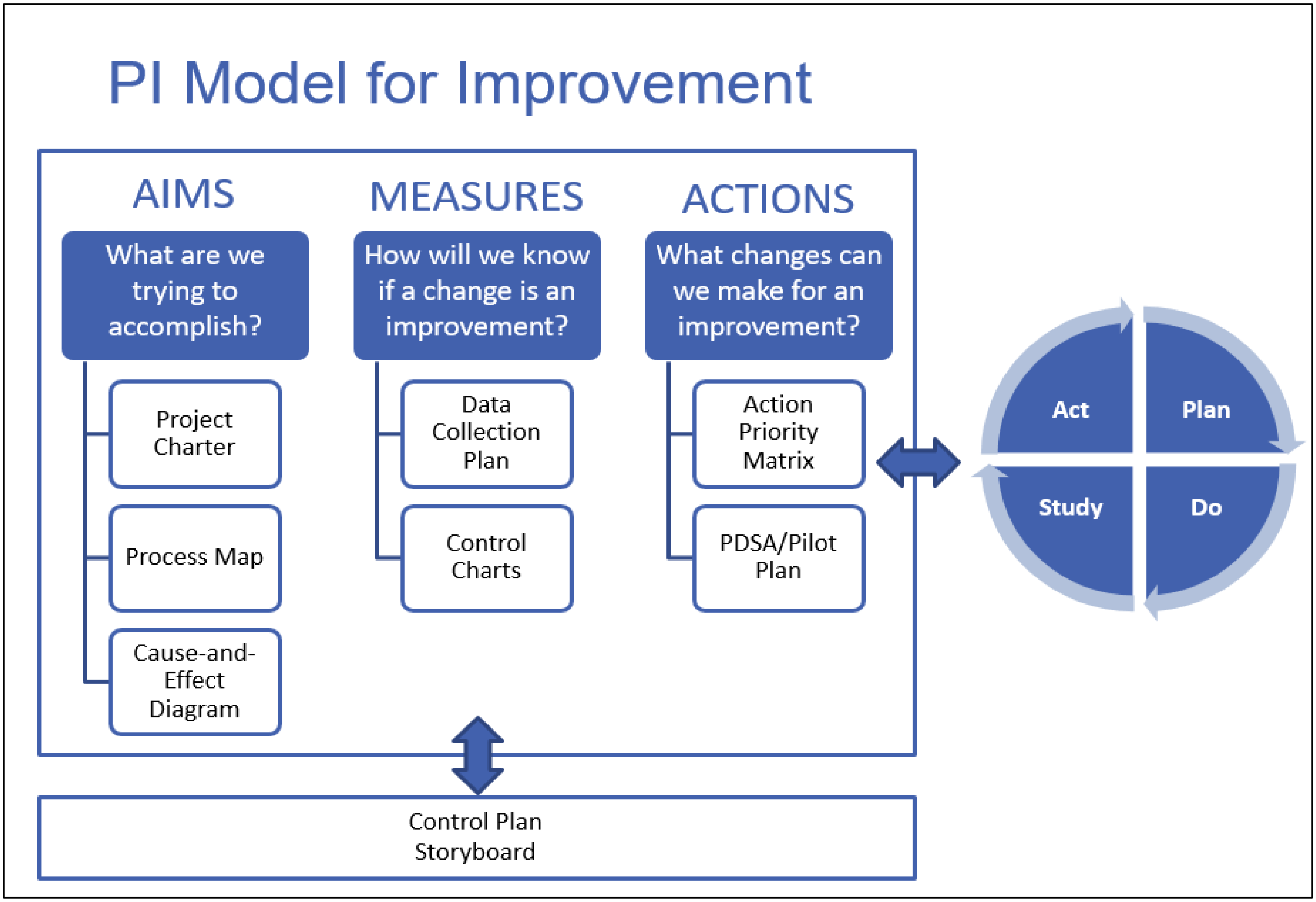

**Supplementary Figure 2: Cause-and-Effect (fishbone) Diagram**

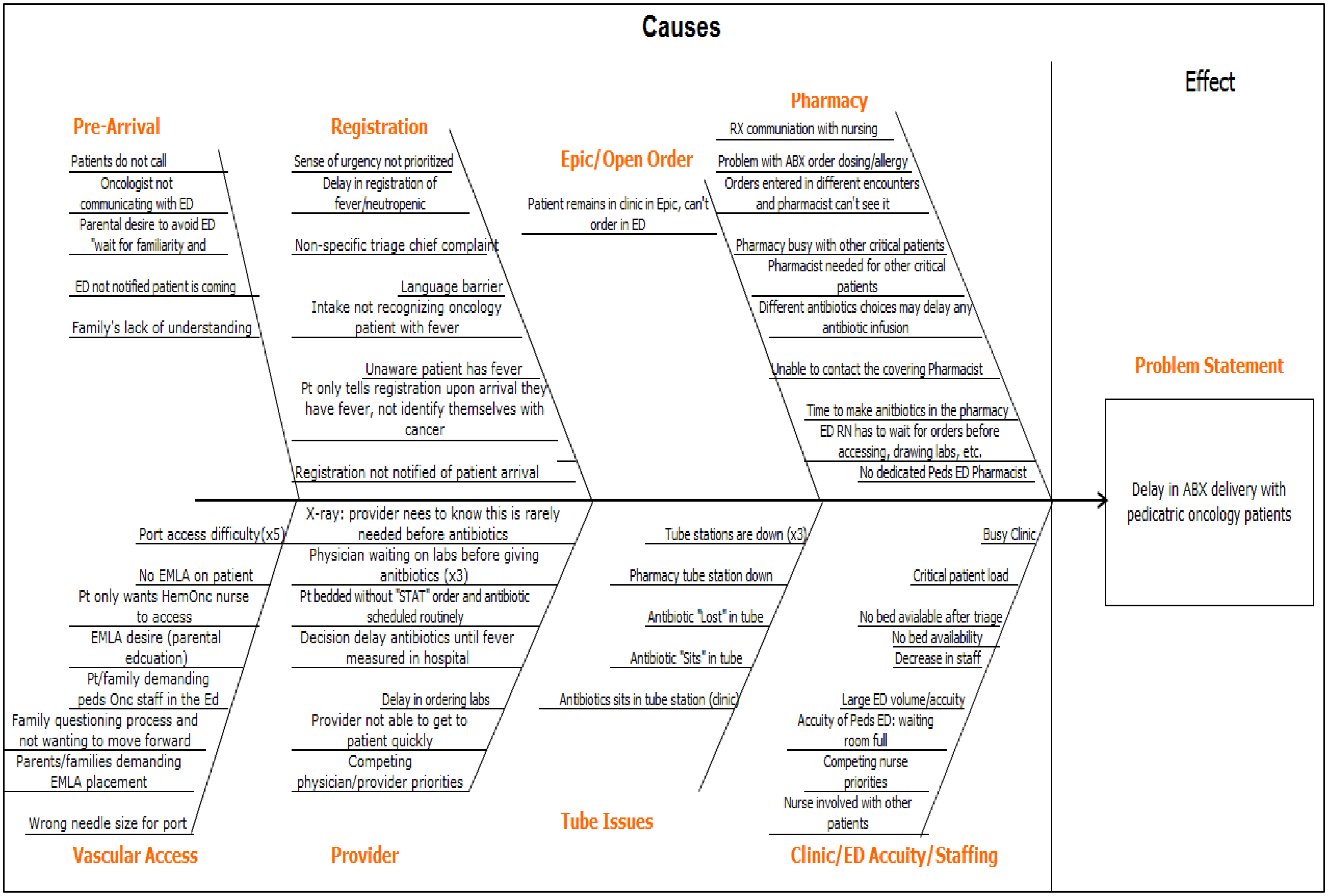

**Supplementary Figure 3: Action Priority Matrix**

High Impact

### Quick-Wins (Low Effort)

- Order 22g in needle for ED
- Notification card for families to present to registration
- Scan Orange Card-place in media tab (with staff education)
- Have a way for pts to identify themselves to ED registration st
- Standardize chief complaints
- Have registration ask specifically if pt is a hem/onc pt(x2)
- Laminated sheet short term-Streamline Antibiotic choice-CTX-Levaquin if allergic to CTX
- Nurse initiated orders that can be started in triage
- Nurse triage protocol for labs-CBC,Blood cultures-ABX-Rocephin if no allergy, Neropenom(?) if allergy
- Any provider call to let pharmacy know if a N/F pt arriving-then we'll be ready and can make sure antibiotic dilution is available
- Notify registration if patient is known to be arriving to ED
- Educate Registration staff about new process and urgency
- Increase education of new oncology patient of ED process
- Patient responsibility card
- New onset canter patients get education at diagnosis about ED care-fast,no EMLA, no H/O nurse
- Clinic: Pt direct telephone number for parents to 9th floor "I hate 716-3364"
- Family orientation session-2 stages
- Be sure note closes prior to ED/inpt admit

### Major Projects (High Effort)

- Order set in EPIC
- Peds specific Pharmacist for Peds ED
- More ED beds and staff
- Specific notifier/color for N/F patients on EPIC trackboard
- Flag for Oncology patients in EPIC
- Create Hem/Onc shock clock button in EPIC to time of register to time of ABX

Low Impact

### Fill-Ins (Low Effort)

- Clinic: When order entered in EPIC. Make sure it is labeled "STAT"
- Staff education on proper freeze spray application when no EMLA
- Quarterly education with ED/Clinic Inpatient nurses on Access
- Clinic: Page/call pharmacy directly not calling the clinical pharmacist 1st
- Pharmacy call the clinic when the tube is sent with ABX
- Obtain Rocephen dilution with longer expiration date from clean room to cut down on prep time
- Make Charge aware of Hem/Onc pt in department
- Fever education and provide thermometer

### Thankless Tasks (High Effort)

- Tube maintenance and reliability

Low Effort — High Effort

**Supplementary Figure 4: Front of the "Fast-Pass" card to signal to ED intake personnel the urgency of this visit**

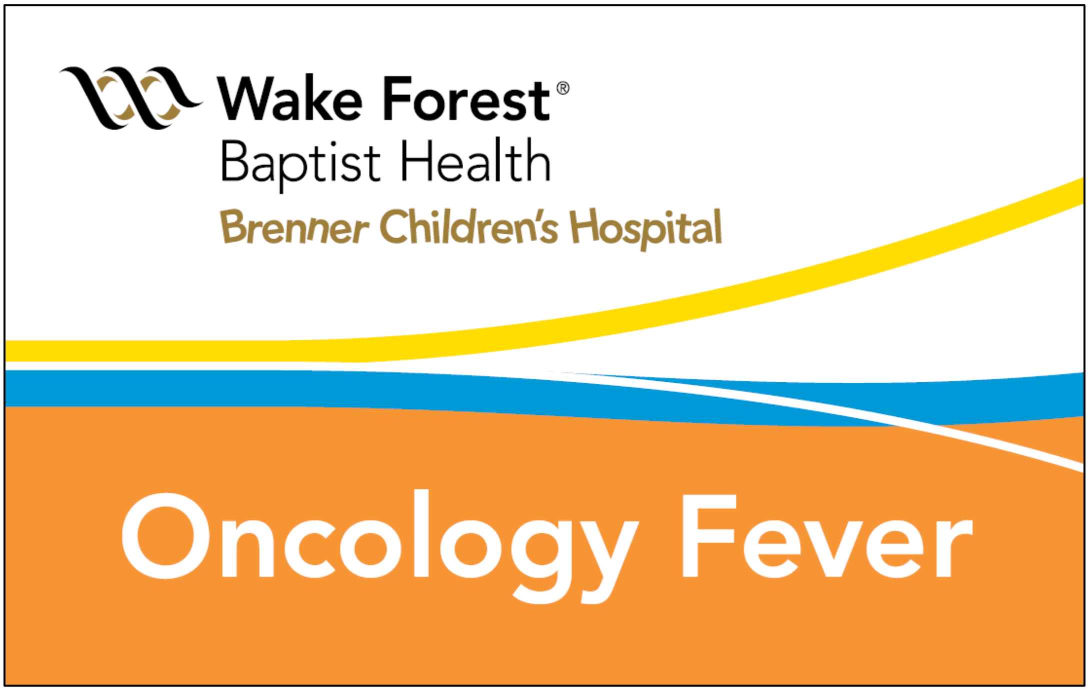

**Supplementary Figure 5: Back of the "Fast-Pass" card to provide information to the ED nursing staff regarding Port-a-Cath needle size and dressing type to be used.**

**Brenner Children's Pediatric Oncology: XXX-XXX-XXXX**

**Initials:** XX **Last Updated**: XX/20XX

**Port Needle Size:** 22g – ¾"

**Dressing Type:** Tegaderm

*Always flush 20mL NS & 5mL hep-lock 10u/mL*

**Supplementary Table 1: Specific times divided into two bins, arrival to EHR order, and EHR order to antibiotic given**

| | **Pre-intervention (October 2016 – June 2017)** | **Post-intervention (July 2017 – June 2019)** | **P value** |
|---|---|---|---|
| *N* | 21 | 360 | |
| Time from arrival to EHR antibiotic order in minutes, *mean* (range); *median* | 20.14 (7-66)<br>17 | 11.40 (0-56)<br>10 | 0.0006**** |
| Time from EHR antibiotic order to antibiotic given in minutes, *mean* (range); *median* | 22.43 (5-65)<br>21 | 19.23 (1-126)<br>16 | 0.0683**** |

****Wilcoxon rank-sum test
